# Exploring [^11^C]CPPC as a CSF1R-targeted PET Imaging Marker for Early Parkinson’s Disease Severity

**DOI:** 10.1101/2023.05.28.23290647

**Authors:** Kelly A. Mills, Yong Du, Jennifer M. Coughlin, Catherine A. Foss, Andrew G. Horti, Katelyn Jenkins, Yana Skorobogatova, Ergi Spiro, Chelsie S. Motley, Robert F. Dannals, Jae-Jin Song, Yu Ree Choi, Javier Redding-Ochoa, Juan Troncoso, Valina L. Dawson, Tae-In Kam, Martin G. Pomper, Ted M. Dawson

**Affiliations:** Department of Neurology, Johns Hopkins University School of Medicine, Baltimore, MD 21287, USA; Johns Hopkins University School of Medicine, Russell H. Morgan Dept. of Radiology and Radiologic Science, Baltimore, MD, USA; Department of Psychiatry and Behavioral Sciences, Johns Hopkins University School of Medicine, Baltimore, MD 21287, USA; Neuroregeneration and Stem Cell Programs, Institute for Cell Engineering, Johns Hopkins University School of Medicine, Baltimore, MD 21205, USA; Department of Pathology (Neuropathology), Johns Hopkins University School of Medicine, Baltimore, MD 21205, USA; Department of Physiology, Johns Hopkins University School of Medicine, Baltimore, MD 21205, USA; Solomon H. Snyder Department of Neuroscience, Johns Hopkins University School of Medicine, Baltimore, MD 21205, USA; Department of Pharmacology and Molecular Sciences, Johns Hopkins University School of Medicine, Baltimore, MD 21287, US; Department of Radiology, University of Texas Southwestern School of Medicine, Dallas, TX, USA (current)

**Keywords:** Parkinson’s disease, microglia, neuroinflammation

## Abstract

Neuroinflammation through enhanced innate immunity is thought play a role in the pathogenesis of Parkinson’s disease (PD). Methods for monitoring neuroinflammation in living patients with PD are currently limited to positron emission tomography (PET) ligands that lack specificity in labeling immune cells in the nervous system. The colony stimulating factor 1 receptor (CSF1R) plays a crucial role in microglial function, an important cellular contributor to the nervous system’s innate immune response. Using immunologic methods, we show that CSF1R in human brain is colocalized with the microglial marker, ionized calcium binding adaptor molecule 1 (Iba1). In PD, CSF1R immunoreactivity is significantly increased in PD across multiple brain regions, with the largest differences in the midbrain versus controls. Autoradiography revealed significantly increased [^3^H]JHU11761 binding in the inferior parietal cortex of PD patients. PET imaging demonstrated that higher [^11^C]CPPC binding in the striatum was associated with greater motor disability in PD. Furthermore, increased [^11^C]CPPC binding in various regions correlated with more severe motor disability and poorer verbal fluency. This study finds that CSF1R expression is elevated in PD and that [^11^C]CPPC-PET imaging of CSF1R is indicative of motor and cognitive impairments in the early stages of the disease. Moreover, the study underscores the significance of CSF1R as a promising biomarker for neuroinflammation in Parkinson’s disease, suggesting its potential use for non-invasive assessment of disease progression and severity, leading to earlier diagnosis and targeted interventions.

**Significance Statement:** This study demonstrates that the Colony Stimulating Factor 1 Receptor (CSF1R) colocalizes with microglial markers in the human brain, and the research establishes elevated CSF1R expression in PD autopsy tissues. Employing [^11^C]CPPC-PET imaging, the study unveils a correlation between increased CSF1R binding and both motor disability and cognitive decline in PD patients. These findings highlight the potential of CSF1R as a novel biomarker for neuroinflammation in PD, offering a non-invasive means to assess disease progression and severity, ultimately contributing to earlier diagnosis and targeted interventions.

## Introduction

The involvement of the innate immune system, including microglial activation of neurotoxic astrocytes(1), is increasingly recognized as playing a pathogenic role in the cascade of events associated with α-synuclein (α-Syn) misfolding and neurodegeneration in Parkinson’s disease (PD). Signaling pathway abnormalities underlying microglial activation to disease-associated microglia converge with those found in genetic forms of PD(2) and neurodegenerative disease more broadly (3, 4).

Based on clinicopathologic analyses of autopsy tissue (5) and the model of fibrillar α-Syn injection in mice (6), the spreading proteinopathy hypothesis holds that fibrillar α-Syn seeds and templates wild type protein, causing toxic intracellular aggregation. Propagation of α-Syn fibrillation and cell to cell spread of pathologic α-Syn is driven, in part, by the innate immune response (7). Once oligomerized, fibrillar α-Syn activates microglia (8, 9), creating a cycle of neuroinflammation and neurodegeneration.

With the interplay between innate immunity and pathogenic α-Syn in the spread of pathologic α-Syn throughout the brain, suppressing this immune activation may allow disease modification in PD (7). A key step in developing targeted therapies to that end is generation of a specific, non-invasive biomarker for microglial activity and proliferation. To date, human *in vivo* assessment of CNS immune activation has primarily involved radiotracers targeting the translocator protein 18 kDa (TSPO)(10), but that approach has limitations. First generation TSPO ligands like [^11^C](*R*)-PK11195 have a poor signal to noise ratio and highly variable kinetic behavior(11). Newer TSPO radiotracers like [^11^C]DPA-713 have higher signal-to-noise but are susceptible to low affinity in persons with a specific TSPO single nucleotide polymorphism(12). More recently, it has also been shown that TSPO expression is not limited to microglia. For example, TSPO expression increases in astrocytes before microglia(13), may be present in neurons(14), and may increase with neuronal activation(15).

As a microglia-selective alternative to TSPO imaging, our team developed a radiotracer that targets colony stimulating factor 1 receptors (CSF1R), a tyrosine kinase involved in proliferation and activation of microglia, with high specificity in the inflamed state(16). While CSF1R may have a role in brain development through expression on specific neuronal populations(17) and microvascular cells(18), in the mature brain, the CSF1R is largely expressed by microglia(17) and is critical for their development and survival(19). The CSF1R radioligand, [^11^C]CPPC, has been shown to have faster kinetics and less off-target cerebellar binding than another leading candidate radiotracer, [^11^C]GW2580, which also shows radiotracer kinetics suboptimal for use in humans due to long scan times(20). [^11^C]CPPC regional volume of distribution (V_T_) values can be estimated in healthy human subjects(21), suggesting its use as a radiotracer in immune-mediated disease states.

Given the need for *in vivo* assessment of microglial density and proliferation at early stages of neurodegeneration in PD to establish causality and potentially, as a biomarker for target engagement in disease modification trials, we studied [^11^C]CPPC in a cohort of persons with early PD and age-similar healthy controls. Our goal was to describe the relationship between PD disease severity and [^11^C]CPPC binding as a cell-type specific marker of microglia.

## Results

### Human postmortem studies

#### CSF1R immunohistochemistry

CSF1R showed colocalization with IBA1+, indicating localization to microglia, in a higher proportion of IBA1+ cells in persons with PD than in controls (Figure 1A & 1B). Homogenized human brain tissue from people with PD showed consistently higher normalized CSF1R expression in the midbrain, cingulate cortex, posterior cingulate cortex, temporal cortex, cerebellar cortex and caudate with the largest differences in the midbrain (p<0.0005), compared to controls with absent neurodegenerative pathology (Figure 1B & D). This work demonstrates that CSF1R is more highly expressed in late stages of PD and that this expression colocalizes with another microglial marker.

**Figure 1.**
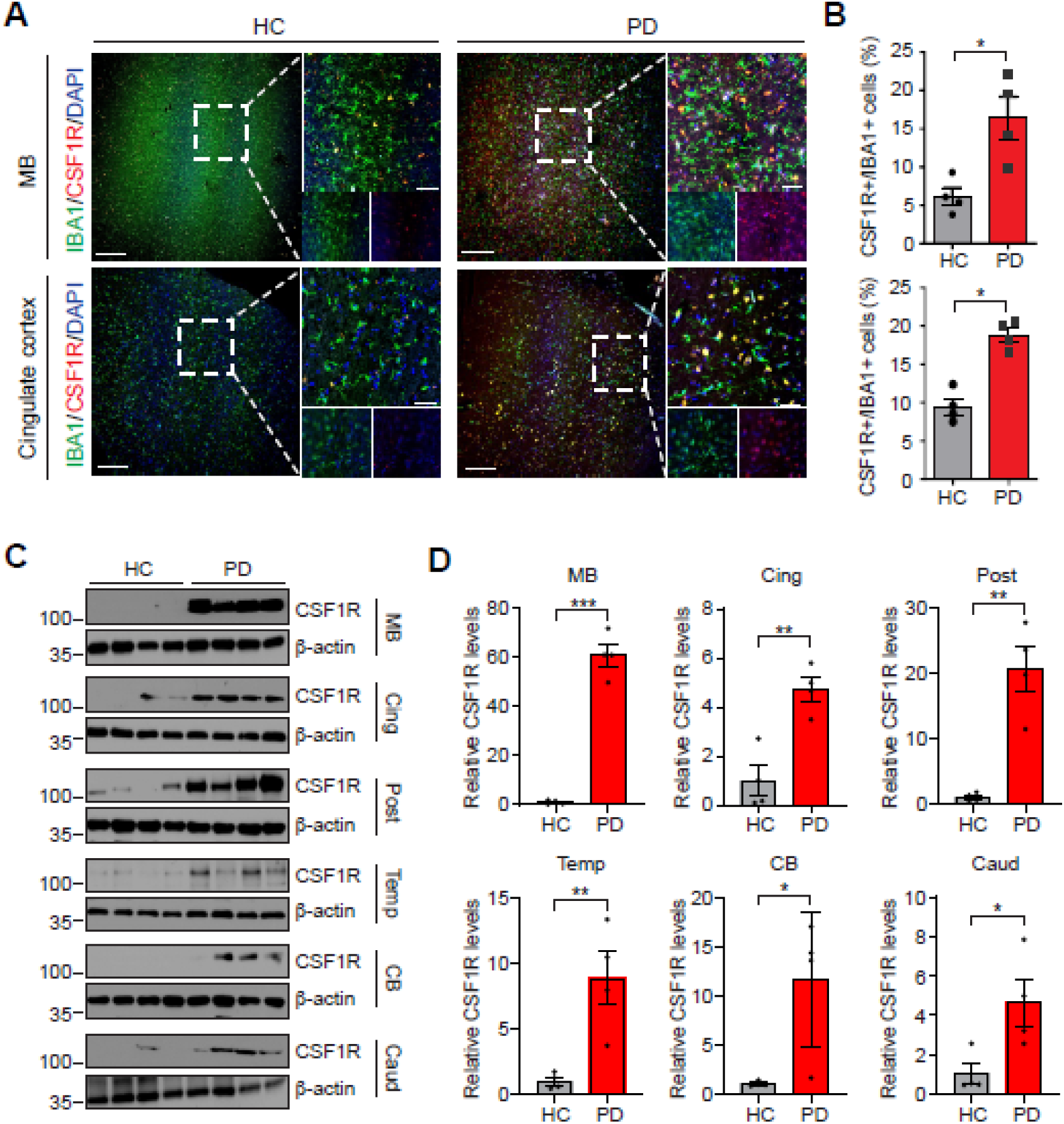
Increased levels of CSF1R in brains of human PD patients. (A) Representative confocal images with DAPI(blue), IBA1(green) and CSF1R(red) in the Midbrain and Cingulate cortex of age-matched healthy control and PD patients. White dashed lines demark the region from where high-magnification images were generated. Scale bars, 200μm (low-magnification images) and 50μm (high-magnification images). (B) CSF1R+ cells with IBA1+ cells are quantified. n = 4 biologically independent sample. Data are shown as the mean ± SEM. p values were determined by unpaired two-tailed Student’s t-tests. *p < 0.05, versus healthy control. (C) Representative immunoblots with CSF1R and β-actin antibodies in the Midbrain (MB), Cingulate cortex (Cing), Posterior cingulate cortex (Post), Temporal cortex (Temp), Cerebellar cortex (CB) and Caudate (Caud) of age-matched healthy control and PD patients. (D) Relative CSF1R levels normalized to β –actin was quantified. n = 4 biologically independent sample. Data are shown as the mean ± SEM. p values were determined by unpaired two-tailed Student’s t-tests. *p < 0.05, **p < 0.005, ***p < 0.0005, versus healthy control.

#### B_max_ measurements

Frozen sections were probed with serially diluted radiotracer with selected cases shown in Figure 2. B_max_ measurements were completed in sections of inferior parietal cortex (IPC), caudate nucleus (CN), midbrain (MB) and basal ganglia (BG) and are displayed in Figure 3 (also, Supplementary Table 1). All values are corrected for a measured 3.65% free (79.35% serum protein bound) radiotracer in the FBS used in these assays. Average grey matter uptake in PD IPC sections was 88.92 ± 6.83 fmol/mg compared with 20.56 ± 10.41 fmol/mg in control. Average white matter uptake in PD IPC was 45.10 ± 1.73 fmol/mg while in control, white matter uptake average is 5.01 ± 2.38 fmol/mg. PD uptake of probe in both regions was about 4-9 times higher than in control. No pattern was observed between male or female uptake. The average GM uptake in PD caudate nucleus was 15.72 fmol/mg while the average GM uptake in control caudate was 6.53 fmol/mg, a 241% increase in binding to PD caudate GM. WM uptake in PD caudate averaged 1.79 fmol/mg while control WM uptake averaged 2.14 fmol/mg. Midbrain samples showed average PD GM uptake as 8.85 fmol/mg while control GM binding averaged 5.73 fmol/mg, a 154% increase in the PD GM sections. WM uptake in PD midbrain averaged 2.68 fmol/mg while control WM uptake in this region averaged 0.51 fmol/mg, nearly 5X lower than PD WM uptake. PD GM uptake in basal ganglia averaged 15.95 fmol/mg while control GM uptake averaged 12.41 fmol/mg, a nearly equal uptake in PD. In WM, PD uptake averaged 5.92 fmol/mg while control WM uptake averaged 1.42 fmol/mg in basal ganglia. There was a statistically significant difference in radiotracer uptake between controls and PD participants in the WM from IPC (ρ = 0.047). uptake.

**Figure 2.**
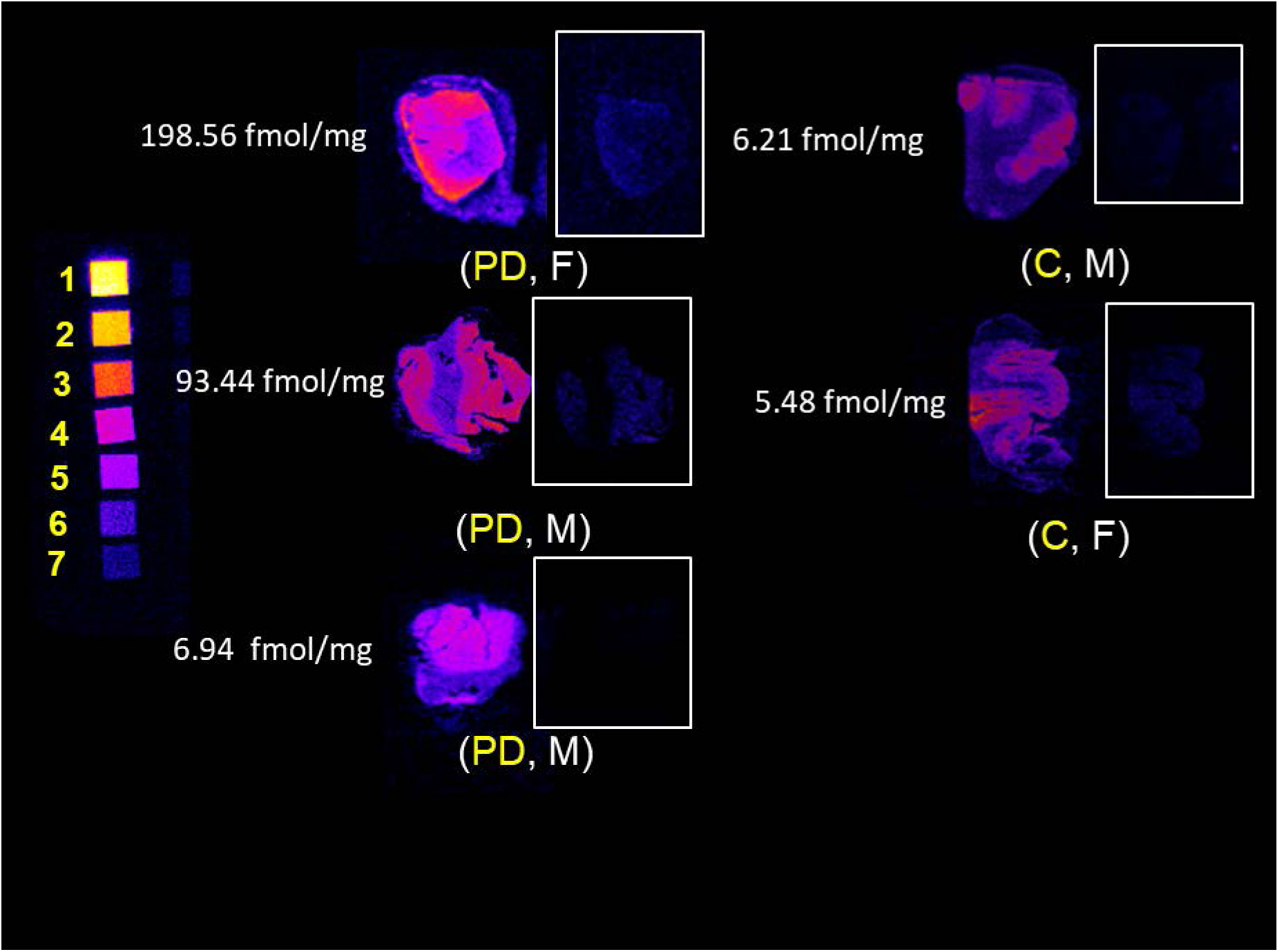
*In vitro* CSF1R autoradiography with ^3^H-JHU11761 in human inferior parietal cortex with and without Parkinson’s disease. Labels show diagnosis of Parkinson’s disease (PD) or C (control) with donor age and sex. Tritium scales standards on the left depict densities beginning at 5.89 nmol/g (1) and serially decrease until (7), 0.09 nmol/g. Grey matter B_max_ is indicated to the left of each case. PD age range 65-72. Control age range 90-97.

**Figure 3.**
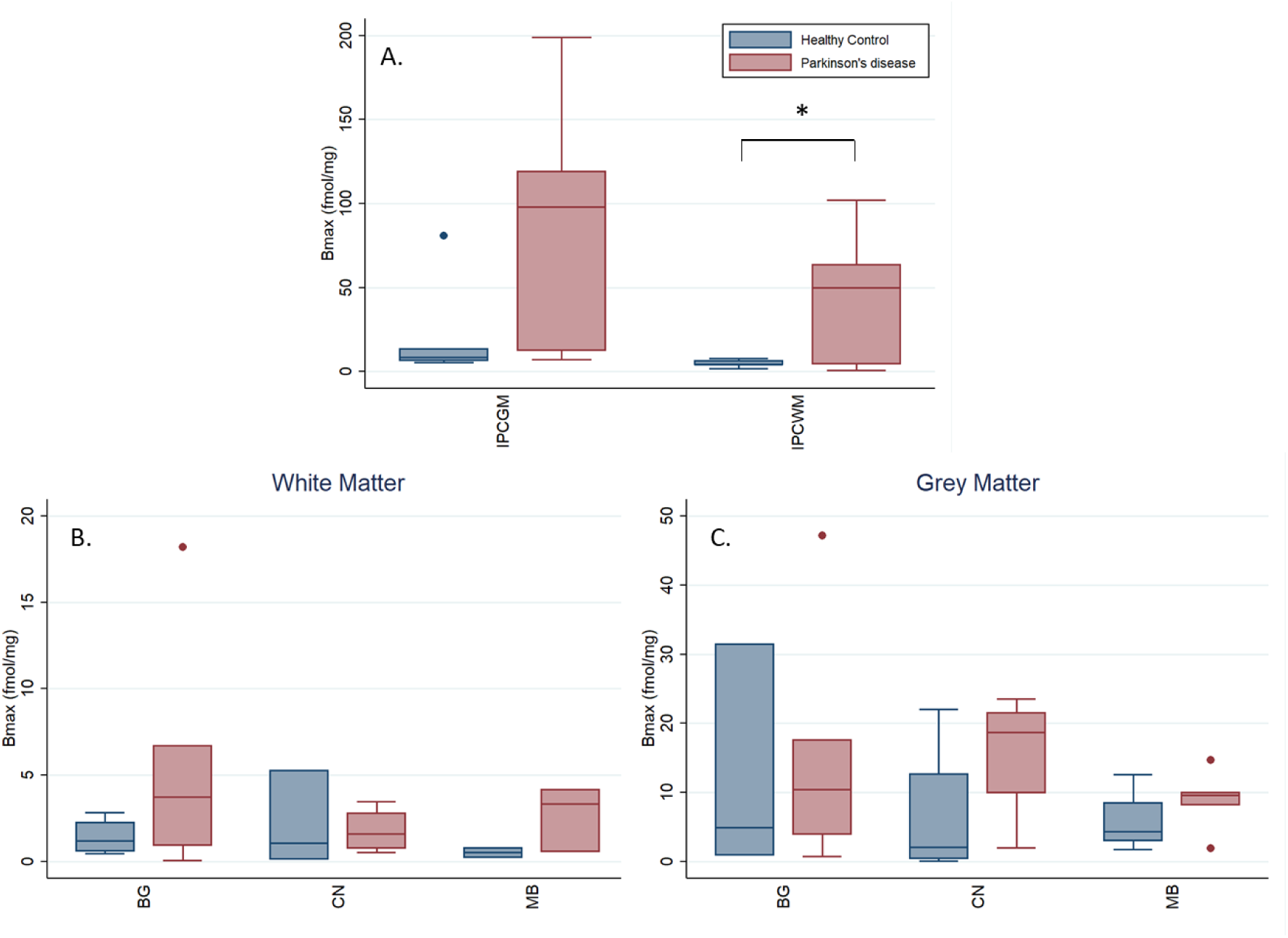
B_max_ of ^3^H-JHU11761 from human frozen sections of A) inferior parietal cortex grey matter (IPCGM) and white matter (IPCWM), and B) white matter and C) grey matter B_max_ in the caudate nucleus (CN), midbrain (MB), and basal ganglia (BG). IPCGM and IPCWM shown separately due to difference in y-axis scale. *p<0.05.

#### Equilibrium binding survey

Supplementary Figure 1 shows the results of a 5 nM binding survey to elicit the number of CSF1R sites in inferior parietal cortex, midbrain and caudate nucleus across four controls and 5 samples with PD. Three controls out of four showed low tracer binding in caudate putamen and midbrain and one PD case also showed low tracer binding across the three subregions. BRC2020 (control) had elevated binding in IPC and caudate nucleus. This may be a consequence of observed cerebrovascular disease and its associated inflammation. Four out of 5 PD cases showed elevated radiotracer binding across gray matter subregions, suggesting elevated CSF1R-related inflammation.

### CSF1R PET Imaging

We consented 12 controls but excluded one due to recent COVID19 infection within the past two weeks and excluded another because of cognitive complaints within domains of memory and executive function. We consented and enrolled twelve participants with PD, all with a Hoehn and Yahr score of 2 or lower and within two years of diagnosis. Of the twelve PD participants, six had PD-MCI based on our criteria. In addition, of the twelve PD participants, eight had “mild” and four had “moderate” disability from motor symptoms based on whether they were equal to or below versus above the median MDS-UPDRS-II score, respectively. Clinical characteristics of these groups are in Table 1.

**Table 1.** Demographics in groups stratified by disease status (PD-all = all participants with clinically established Parkinson’s disease (PD)) or disease severity. PD-NC = PD with normal cognition; PD-MCI = PD with mild cognitive impairment by level 1 criteria; PD-Mild = PD with ≤ median score for cohort on MDS-UPDRS Part II; PD-Moderate = PD with > median score for cohort on MDS-UPDRS Part II. MoCA = Montral Cognitive Assessment, BDI-2 = Beck Depression Inventory-2, MDS-UPDRS = Movement Disorder Society Unified Parkinson’s Disease Rating Scale

|  | Controls |  | PD-all |  | PD-NC |  | PD-MCI |  | PD-Mild |  | PD-Moderate |  |
| --- | --- | --- | --- | --- | --- | --- | --- | --- | --- | --- | --- | --- |
| N | 10 |  | 12 |  | 6 |  | 6 |  | 8 |  | 4 |  |
|  | μ | SD | μ | SD | μ | SD | μ | SD | μ | SD | μ | SD |
| Age (yr) | 65.3 | 4.4 | 68.8 | 5.9 | 68.2 | 6.9 | 67.4 | 6.5 | 67.2 | 6.6 | 71.9 | 2.0 |
| Levodopa (mg/day) | 3 | 2 | 336 | 185 | 440 | 152 | 22 | 5 | 354 | 133 | 300 | 300 |
| MDS-UPDRS III | 0.2 | 0.4 | 24.7 | 8.0 | 21.4 | 5.5 | 6.0 | 4.3 | 22.0 | 2.5 | 30.0 | 12.7 |
| MDS-UPDRS II | 26.7 | 2.8 | 6.1 | 4.9 | 6.2 | 4.8 | 27.8 | 1.2 | 3.1 | 2.0 | 12.0 | 3.2 |
| MoCA | 42.2 | 8.9 | 25.2 | 3.3 | 28.0 | 1.2 | 46.7 | 12.8 | 25.5 | 2.6 | 24.5 | 4.8 |
| BDI-2 | 1.9 | 2.5 | 6.5 | 4.4 | 7.0 | 4.2 | 7.0 | 3.7 | 5.8 | 4.6 | 8.0 | 4.2 |

The mean [^11^C]CPPC V_T_ was computed using the Logan method and plotted across ROIs in healthy controls and those with both mild– or moderate-PD based on disability from motor symptoms quantified by the MDS-UPDRS part II (Figure 4). ANOVA testing for the difference in mean V_T_ between these groups showed significant difference by group in the brainstem, cerebellar cortex, striatum, frontal cortex, hippocampus, pallidum, thalamus, and frontal, parietal, and temporal cortices, though the difference between groups was only statistically significant in the striatum when adjusting for multiple comparisons (p<0.004) (Supplementary Table 2). Tukey’s post hoc testing showed that this difference was driven by the differences between the moderate motor disability and mild motor disability groups in the brainstem, cerebellar cortex, striatum, pallidum, hippocampus, thalamus, and frontal, parietal, and temporal cortices and between moderate motor disability and control groups in the cerebellar cortex, striatum, hippocampus, thalamus, and frontal and parietal cortices. No regions showed a statistically significant difference between healthy controls and mild motor disability groups in a group-wise comparison but V_T_ in multiple regions showed a positive correlation with motor-related disability. Greater motor disability was associated with higher [^11^C]CPPC binding in the brainstem, cerebellar cortex, thalamus, hippocampus, striatum, pallidum, and temporal, occipital, frontal, posterior cingulate, and parietal cortices, though only the brainstem (r = 0.78, p = 0.003) and temporal cortex (r = 0.78, p = 0.003) remained statistically significant after adjusting for multiple comparisons (Table 2). Representative scatter plots from the brainstem and temporal cortex, fit with a linear representation of this correlation, are seen in Figure 5.

**Figure 4.**
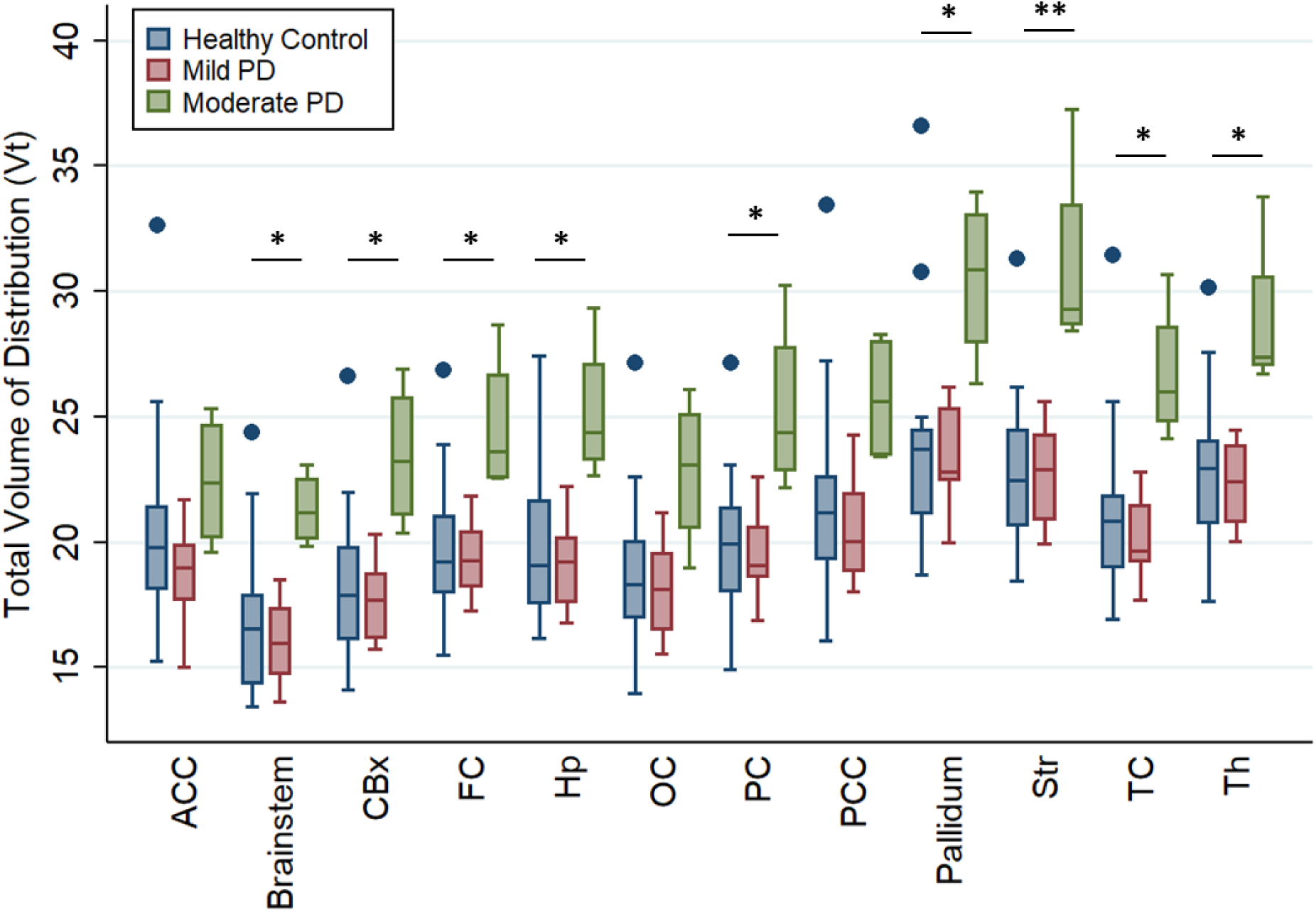
Regional total volume of distribution (V_T_) of [^11^C]CPPC, a CSF1R PET radioligand, in people with mild or moderate PD, defined by motor disability, and controls of a similar age. * = p<0.05 and ** = p<0.004 for ANOVA and post-hoc test between Moderate PD vs. Mild PD and or Moderate PD vs. healthy controls. ACC = anterior cingulate cortex, CBx = cerebellar cortex, FC = frontal cortex, Hp = hippocampus, OC = occipital cortex, PC = parietal cortex, PCC = posterior cingulate cortex, Str = striatum, TC = temporal cortex, Th = thalamus. Dot = outliers.

**Figure 5.**
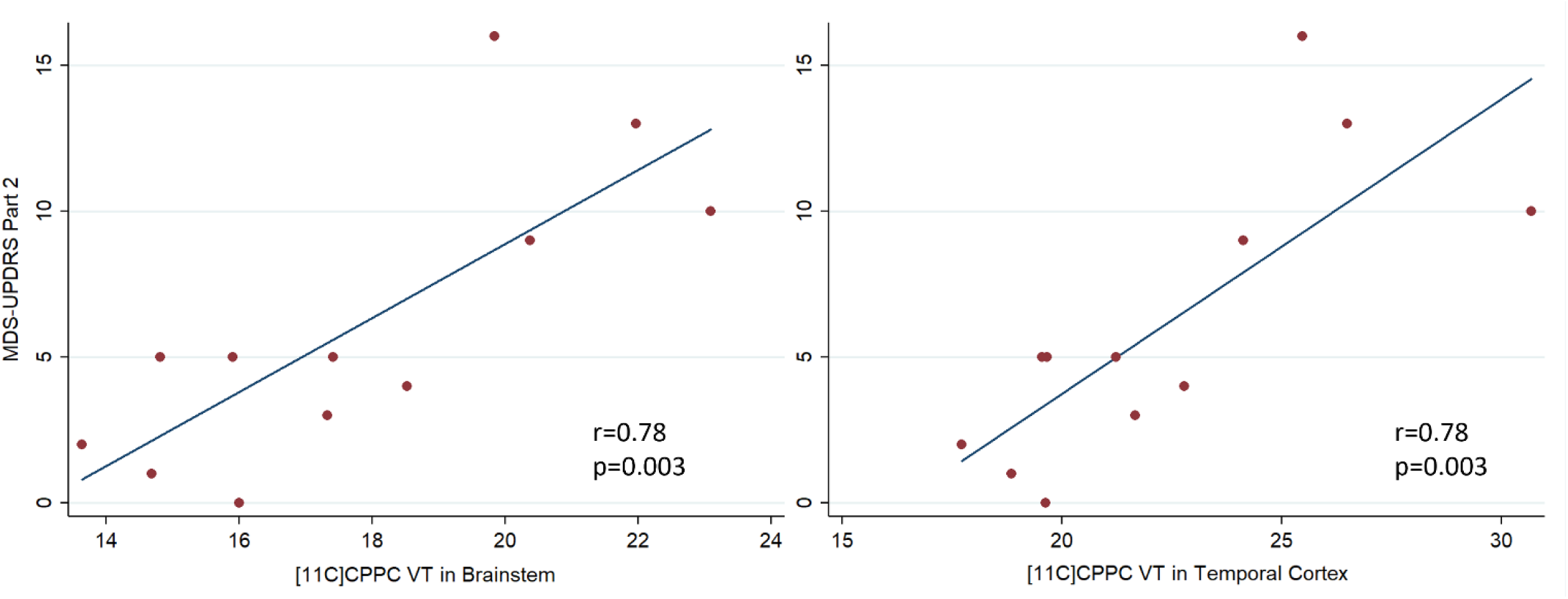
Scatter plots and linear relationship between [^11^C]CPPC V_T_ and ADL disability from motor symptoms, measured by MDS-UPDRS Part II in regions of interest that showed a statistically significant relationship (p<0.005).

**Table 2.** Results of Pearson correlation between MDS-UPDRS Part 2 and regional [^11^C]CPPC V_T_ in 12 persons with Parkinson’s disease. *p<0.05 (unadjusted), **p<0.005 (with Bonferroni correction).

| Region | r | p |  |
| --- | --- | --- | --- |
| Anterior cingulate cortex | 0.488 | 0.107 |  |
| Brainstem | 0.776 | 0.003 | ** |
| Cerebellar cortex | 0.7422 | 0.006 | * |
| Frontal cortex | 0.672 | 0.017 | * |
| Hippocampus | 0.656 | 0.021 | * |
| Occipital cortex | 0.727 | 0.007 | * |
| Parietal cortex | 0.692 | 0.013 | * |
| Posterior cingulate cortex | 0.692 | 0.013 | * |
| Pallidum | 0.629 | 0.028 | * |
| Striatum | 0.668 | 0.014 | * |
| Temporal cortex | 0.775 | 0.003 | ** |
| Thalamus | 0.687 | 0.014 | * |

Participants with PD were also stratified by cognitive status, and at a group level, [^11^C]CPPC was not higher in any region in participants with PD-MCI than those with PD-NC or healthy controls (ANOVA p>0.05 for all regions). Pearson correlation between regional [^11^C]CPPC V_T_ and a measure of global cognitive function, MoCA, were of moderate strength at best (putamen; r = –0.42, p = 0.18). Because this cohort has early PD, there was a ceiling effect of the MoCA with 4 of 5 PD-NC patients having a 28 or 29 out of 30 total possible points. However, worse phonemic verbal fluency was associated with higher [^11^C]CPPC V_T_ in multiple regions of interest (Table 3), but statistical testing of these correlations did not survive correction for multiple comparisons with alpha = 0.004.

**Table 3.** Results of Pearson correlations between phonemic verbal fluency regional [^11^C]CPPC V_T_ in 12 persons with PD, adjusted for age.

| Region | $\beta$ | p | |
| --- | --- | --- | --- |
| Brainstem | -0.70 | 0.012 | * |
| Cerebellar Cortex | -0.70 | 0.011 | * |
| Thalamus | 0.63 | 0.027 | * |
| Hippocampus | -0.59 | 0.045 | * |
| Temporal cortex | -0.69 | 0.014 | * |
| Occipital cortex | -0.74 | 0.006 | * |
| Frontal cortex | -0.59 | 0.044 | * |
| Parietal cortex | -0.64 | 0.025 | * |
| Caudate | -0.41 | 0.188 |  |
| Putamen | -0.70 | 0.011 | * |
| Pallidum | -0.69 | 0.013 | * |
| Anterior cingulate | -0.35 | 0.266 |  |
| Posterior cingulate | -0.63 | 0.027 | * |

## Discussion

In response to the substantial heterogeneity in clinical presentation and pathogenic mechanisms in PD(22), precision medicine strategies are being sought to identify mechanistic subtypes of PD and target that subtype with a relevant disease-modifying strategy. Especially in sporadic PD, an increasing focus has been placed on the role of innate CNS immunity(23), with evidence of elevated innate immune activity in at least a subset of PD patients(24). This study shows that in human brain, CSF1R co-localizes with microglial markers and is elevated in autopsy tissue from PD patients compared to controls, and presents data on the use of [^11^C]CPPC to measure CSF1R binding in early stage PD patients. This study adds to the growing literature on timing of microglial response in neurodegenerative disease by showing correlations between microglial proliferation and motor and cognitive disease severity in PD even at an early stage and that this can be measured with a novel radioligand targeting CSF1R, which has advantages over previously used PET ligands to detect inflammation in PD.

These PET data are an advance from previous work that showed elevation of TSPO in PD participants versus those without known neurodegeneration using PET. Several issues complicate the use of TSPO ligands in research and as potential biomarkers of innate immunity: Though most highly expressed in activated microglia, TSPO may be increased in expression by activated astrocytes even before increased TSPO expression by microglia in neurodegeneration.(13) TSPO may also be found in neurons where it increases with neuronal activation(15). While initial studies with [^11^C]-PK11195 showed higher binding in PD versus controls(25), a subsequent study showed overlap between PD and healthy controls and that treatment with an anti-inflammatory unexpectedly *increased* binding potential in the same brain regions(26). While not susceptible to genetic variation in TSPO binding affinity like later generation TSPO ligands, [^11^C]-PK11195 also has a low signal-to-noise ratio due to off-target binding(27). Later generation TSPO ligands like [^18^F]DPA-714 have higher affinity but this affinity can be substantially reduced by a SNP in the TSPO gene. In a large study comparing [^18^F]DPA-714 binding in people with PD and controls, even after persons with low affinity were excluded based on genotyping, significant differences in binding between PD and controls were only found in the 40 high-affinity binders, but not in the 27 mixed-affinity binders(24). This indicates that use of new generation TSPO ligands to diagnose those with elevated innate immunity or to track microglial activation after an intervention would be limited to high-affinity binders, which are expected to comprise only 49% of the population(12). In our older, healthy controls, and in the least-affected PD group, the variance in binding was relatively small without outliers to suggest high natural variance in binding. Thus, our finding of a potentially more specific marker of microglial activation that is not known to have genetic variation in binding affinity for its target, represents an advance with a more specific, reliable marker of the microglial proliferation and/or activation in PD.

While this study cannot establish a causal link between microglial activation and α-synuclein-related neurodegeneration, it aligns with preclinical and neuropathological evidence that supports a contributory role in cell death. Microgliosis precedes cell death in α-synuclein over-expression animal models (28) and in human autopsy studies(23) and at least two mechanisms have been proposed: 1) direct or indirect (via activation of neurotoxic astrocytes) toxicity to neurons by α-synuclein-activated microglia (1, 23, 29) and 2) microglia-mediated enhancement of α-synuclein oligomerization(30) and spread throughout the brain(31, 32). Since some of the most promising disease-modifying agents in PD(33) are thought to act via microglia(7), having an *in vivo* biomarker for microglial activity may facilitate therapy development and hasten drug trials as an indicator of target engagement in proof-of-principle studies and clinical trials.

In conclusion, these data suggest that using [^11^C]CPPC as a radioligand of CSF1R can detect microgliosis in early PD with at least moderate severity in patients, and that variance in [^11^C]CPPC binding within patients with PD correlates with disability from motor dysfunction and cognitive impairment. [^11^C]CPPC could be useful for identifying potential participants with microgliosis for trials that target immune activation in PD or for evaluating target engagements in such studies.

## Materials and Methods

### Human postmortem brain tissues

This research uses anonymous autopsy material; the frozen tissues and formalin-fixed sections of each brain regions from subjects with PD (n=5) and age-matched controls (n=4) were obtained from the Brain Resource Center, Division of Neuropathology, Department of Pathology of Johns Hopkins University School of Medicine.

### Immunohistochemistry

Immunohistochemistry (IHC) was performed on human brain sections including cingulate cortex and midbrain. Primary antibodies and working dilutions used were as follows: CSF1R (NBP2-37292, 1:100) and IBA1 (Wako 019-19741, 1:500). Brain sections were deparaffinized with 4 changes of xylene for 2 minutes each and hydrate in 4 changes of 100% ethanol for 2 minutes each and 4 changes of 95% 1 minute each. After washing for 5 minutes, sections were incubated in peroxidase blocking solution, followed by being blocked with 4% donkey serum (Sigma-Aldrich)/ PBS plus 0.2% Triton X-100 and incubated with primary antibodies. After brief washes with PBS containing 0.2% Triton X-100, brain sections were incubated with corresponding secondary antibodies conjugated with fluorescent dyes (Alexa Fluor 488–conjugated donkey antibody to rabbit IgG for IBA1, Alexa Fluor 568–conjugated donkey antibody to mouse IgG for CSF1R). The images were acquired by confocal scanning microscopy (LSM710, Carl Zeiss), followed by processed by the Zen software (Carl Zeiss).

### Western blot analysis

Human postmortem brain tissues were homogenized and prepared in lysis buffer [50 mM Tris-HCl (pH 7.4), 150 mM NaCl, 1 mM EDTA, 1% Triton x-100, 0.5% SDS, 0.5% sodium-deoxycholate, phosphatase inhibitor mixture I and II (Sigma-Aldrich, St. Louis, MO), and complete protease inhibitor mixture (Roche, Indianapolis, IN)]. The homogenates were rotated at 4°C for 30 min for complete lysis, and centrifuged at 15,000 x g for 20 min. The supernatants were used for protein quantification using the BCA assay (Pierce, Rockford, IL). Samples were separated using SDS-polyacrylamide gels and transferred onto nitrocellulose membranes. The membranes were blocked with 5% non-fat milk in TBS-T (Tris buffered saline with 0.1% Tween-20) for 1 h, probed using CSF1R antibody (Invitrogen, 14-1152-82), followed by incubation with the HRP-conjugated secondary antibodies (Cell signaling). The bands were visualized by ECL substrate.

### Autoradiography

#### B_max_ measurements

Unfixed, frozen human brain tissues were obtained from the JHU Brain Resource Center from deaths occurring from PD or non-Parkinson’s/non-Alzheimer’s controls. Human autopsy tissues were cryosectioned to 16 µm thickness on charged glass slides and kept frozen at –80°C until use. Serially diluted solutions containing [^3^H]-JHU11761 (4-cyano-N-(4-(4-[3H]methylpiperazin-1-yl)-2-(4-methylpiperidin-1-yl)phenyl)-1H-pyrrole-2-carboxamide) in FBS (Sigma-Aldrich, St. Louis, MO) spanned 9 nM through 9 pM. Each solution was applied to cognate slides and allowed to bind at ambient temperature for 10 min. Solutions were then rapidly aspirated off and the slides washed in ice cold PBS (St. Louis) for 2 min. The PBS was aspirated off and any remaining moisture removed. The dry slides were then loaded into a Hypercassette (Amersham RPN-11647) and exposed to a charged phosphorscreen (Cytiva, BAS-TR 2040) for 5 days. The screen was scanned using a Typhoon 9500IP Phosphorimager (Molecular Dynamics) and data visualized and quantitated using ImageJ software (Sourceforge.net). ROIs were drawn over grey and then white matter to quantitate radioligand uptake and then transformed with the tritium scales standard (ARC, St. Louis, MO). Values were graphed using Prism software (GraphPad, Boston, MA) to fit a Scatchard plot where the saturation binding value (B_max_) was obtained for each case and then as average PD and control values. Plasma protein binding of 3H-JHU11761 in lot-specific FBS was determined using 6 nM radioligand binding at ambient temperature followed by loading a 0.5 mL aliquot of FBS into an Amicon Ultra-0.5 mL 30K purification device, which was centrifuged at 14,000 x g for 30 min at room temperature. The filtrate was then saved and the remaining supernatant was recovered by centrifugation at 1,000 x g for 2 min. The filtrate, recovered supernatant and filter itself were primed with ScintiVerse scintillation cocktail and were counted separately in a Beckman Coulter LS 6500 (Brea, CA) multi-purpose scintillation counter. The filtrate represented “free” or unbound radiotracer while the supernatant represented protein bound radiotracer (≥30 kDa protein-bound). The filter device represented non-specific binding and this value was subtracted from both supernatant and filtrate values. Percent free radiotracer was then used as a correction factor for measured Bmax values. A two-sided, two sample t-test was applied to Bmax averages between AD and control groups for each subregion as well as gray and white matter.

#### Equilibrium binding survey

Basal ganglia (containing caudate-putamen), midbrain and inferior parietal cortex containing slides (n = 3 and n= 1 blocking for each region and each case: four control, 5 PD) were obtained along with the B_max_ cohort and have the same properties. They were probed with 5 nM [^3^H]-JHU11761 in FBS or along with 50 µM unlabeled compound (blocking) in FBS for 10 min at ambient temperature. Binding solution was aspirated and the tissues were washed with ice cold PBS for 2 min. The PBS was then aspirated off and the slides dried before loading into a Hypercassette for exposure to a charged phosphorscreen. Exposures took 5 days and were processed identically to the slides for B_max_ measurements although quantitation involved only ROI drawing and comparison with the standard curve obtained from the tritium microscales.

### PET Imaging Subjects

We enrolled persons between ages 50-80 with a clinically established Parkinson’s disease(34) within 2 years of diagnosis and with Hoehn and Yahr stage 2 or lower. People in this state of disease are fully ambulatory and mostly independent. Healthy subjects age 50-80 were recruited from PD and PD caregiver support groups, families of persons with PD, or the general public. We excluded potential participants with other systemic autoimmune disorders, recent infection, anti-inflammatory or immunosuppressant use, a history of moderate or severe TBI, or other known neurodegenerative diseases. Safety labs, including chemistry panel, liver function tests, serum pregnancy test (if female and premenopausal), and EKG were performed and patients were excluded for lab values >1.5 times the upper limit of normal. The study was performed with approval by the Johns Hopkins Institutional Review Board and was compliant with the Declaration of Helsinki. Patients underwent informed consent prior to participation in the study protocol.

At the baseline visit, all participants underwent examination for parkinsonism and if absent, the Prodromal PD Calculator was administered and healthy controls were only enrolled as such if the likelihood ratio for prodromal PD did not meet or exceed that which would provide 80% likelihood of eventual PD, stratified by age(35, 36), and if they did not meet criteria for clinically established Parkinson’s disease(34). Participants with a prior diagnosis of PD were examined for parkinsonism and if present, the Movement Disorder Society Clinical Diagnostic Criteria for Parkinson’s Disease were applied. Those meeting criteria for clinically established Parkinson’s disease were enrolled in the “Parkinson’s disease” cohort.

### Clinical Assessments

All participants underwent the Movement Disorders Society-Unified Parkinson’s Disease Rating Scale (MDS-UPDRS) parts I-IV in the medicated state. To explore the relationship between disease severity and CSF1R imaging, participants were classified as “mild PD” if their MDS-UPDRS Part II was at the median (5) or lower, or “moderate PD” if above the median. Part II was used because it is more sensitive to disease progression across the entire spectrum of disease severity, including early disease(37). Other assessments included Montreal Cognitive Assessment (MoCA), phonemic (letter) verbal fluency, Beck Depression Rating Scale-2, University of Pennsylvania Smell Identification Test, in addition to a medical history and medication history. Participants with PD were classified as having mild cognitive impairment (PD-MCI) by level 1 criteria(38) if they had a subjective report of cognitive impairment (MDS-UPDRS part 1, question 1 ≥1) and an abnormal measure of global cognitive impairment (MoCA<26).

### MRI

Participants received a brain MRI for PET co-registration and to screen for anatomic variations or lesions that would preclude analysis of PET data at a group level. MRI’s were performed on a Philips dStream Ingenia Elition 3T with a 3D-acquired T1-wieghted sequence, T2 sequence, and iron-sensitive sequences. FreeSurfer(39) image analysis suite was used for automated segmentation of MRI into regions of interest (ROIs) in PD: brainstem, cerebellum, thalamus, hippocampus, temporal cortex, occipital cortex, frontal cortex, parietal cortex, pallidum, striatum, anterior cingulate cortex, and posterior cingulate cortex.

### PET Imaging

The synthesis of [^11^C]CPPC occurred in the Johns Hopkins PET Center, where radiochemical purity was confirmed at >95%(16). The target injection radioactivity was 740 MBq, with a max of 777 MBq and minimum of 703 MBq. Participants were fitted with a thermoplastic face mask to minimize head motion. A radial arterial catheter was inserted for serial arterial blood sampling and an intravenous catheter was inserted for PET radiotracer injection. A 128-slice low-dose CT was administered for attenuation correction before the injection. A 90-minute emission scan on a Siemens Biograph mCT PET/CT began with IV injection of 740 MBq of [^11^C]CPPC. Serial arterial blood was at first drawn as fast as possible for 90 seconds, then at increasingly longer intervals between blood samples thereafter. Select samples at 5, 10, 20, 30, 60, and 90 minutes were analyzed with high performance liquid chromatography (HPLC) for radioactive plasma metabolites (21). Thirty frames (four of 15 seconds, four of 30 seconds, three of 1 minute, two of 2 minutes, 5 of 4 minutes, and twelve of 5 minutes) from the 90 minutes emission scan were reconstructed using the iterative ordered subset expectations maximization algorithm and corrected for attenuation, randoms, scatter, and time-of-flight. PET data processing and tracer kinetic modeling were performed as previously described(21) using PMOD (v3.7, PMOD Technologies Ltd., Zurich, Switzerland). Briefly, inter-frame motion correction was first applied by frame-by-frame matching to a reference frame generated from the average of the frames from 30-60 minutes post-injection. Next, the PET data were transformed to MRI space through rigid registration. The MRI segmented ROIs were then applied to the registered PET images to generate a time activity curve for each region. The total distribution volume V_T_, which reflects the equilibrium ratio of [^11^C]CPPC concentration in tissue to the concentration in arterial plasma was finally calculated using Logan graphical analysis(40) with metabolite corrected plasma input function.

### Statistical analyses

For immunohistochemistry, the mean percentage of cells in each sample with CSF1R+/IBA1+ staining were compared with unpaired two-tailed Student’s t-tests for each region. For immunoblotting results, CSF1R levels were reported as the mean CSF1R level normalized to β-actin and compared between PD and control groups using a two-tailed Student’s t-test. For human PET imaging, Pearson correlation was performed between age and clinical outcome variables including MDS-UPDRS Part II, MoCA, and verbal fluency and since these were not significant, future associations between clinical and imaging variables were unadjusted. Independent samples t-tests were used to evaluate the difference in V_T_ between healthy controls and participants with PD in each ROI. Given a high degree of heterogeneity in regional V_T_ in the PD group, for each region of interest, [^11^C]CPPC V_T_ was correlated with disease severity measured by the MDS-UPDRS Part II, the most clinically relevant measure of disease severity (37). Based on findings from this analysis, PD participants were stratified in the in the domain of disability from motor symptoms into “mild” disease (≤ median MDS-UPDRS Part II score of 5) or “moderate” disease (>median MDS-UPDRS Part II score of 5). ANOVA was performed for each region of interest to test for any difference in variance of V_T_ between 3 groups: healthy controls, “mild” PD, or “moderate” PD. To further explore the heterogeneity of [^11^C]CPPC V_T_ in PD participants as it relates to variance in the cognitive domain, regional V_T_ was assessed for correlation with verbal fluency, a cognitive function known to involve multiple brain regions and that is affected early in PD(41).

Analyses were adjusted for multiple comparisons of 13 ROI’s using Bonferroni correction, with α set at 0.004. Given that this is a first-in-PD pilot study using [11C]CPPC PET to image the CSF1R, the statistical significance of unadjusted results were also presented.

## Data Availability

All data produced in the present study are available upon reasonable request to the authors at the end of the performance period for funding.

## Acknowledgments

The authors would like to thank the PET Radiotracer Center staff (Andrew Hall, Hong Fan, Daniel P. Holt, and William B. Mathews) for their expertise in the synthesis of the radiotracer used in this study and the PET Center staff (Karen Edmonds) for her scanning expertise. PET imaging was funded by the Michael J. Fox Foundation and the RMS Family Foundation. Radiotracer development was funded by the NIH (R01AG066464). Postmortem brain tissues were provided by the Brain Resource Center supported by the Johns Hopkins Alzheimer’s Disease Research Center NIH P30 AG066507 and BIOCARD study NIH U19 AG033655.

