## Supplementary Table 1, Supplementary Table 2, Supplementary Figure 1 for "Exploring [^11^C]CPPC as a CSF1R-targeted PET Imaging Marker for Early Parkinson’s Disease Severity"

Supplementary Data

Supplementary table 1.

|  |  |  | Inferior parietal cortex | | Caudate | | Midbrain | | Basal ganglia | |
| --- | --- | --- | --- | --- | --- | --- | --- | --- | --- | --- |
| Group | Sex | Age Range | GM | WM | GM | WM | GM | WM | GM | WM |
| Control | F | 90-94 | 5.48 | ND | 0.71 | ND | 4.40 | ND | 0.85 | 0.47 |
| Control | M | 70-74 | 8.79 | 7.99 | ND | ND | ND | ND | ND | ND |
| Control | F | 65-79 | 80.74 | 4.19 | ND | ND | ND | ND | ND | ND |
| Control | M | 80-84 | 13.58 | 6.68 | 21.99 | 5.27 | 12.60 | ND | 31.50 | 2.84 |
| Control | F | 60-69 | 8.34 | 1.86 | 3.39 | 1.04 | 1.74 | 0.22 | 4.87 | 0.72 |
| Control | M | 95-99 | 6.43 | 4.34 | 0.02 | 0.10 | 4.21 | 0.79 | ND | 1.66 |
| PD | M | 75-79 | 119.14 | 63.70 | 1.93 | 0.52 | 1.88 | 0.56 | 0.72 | 0.05 |
| PD | M | 90-94 | 12.64 | 4.39 | 17.95 | 2.15 | 14.63 | 4.15 | 17.62 | 6.71 |
| PD | M | 70-74 | 7.26 | 0.74 | ND | ND | 8.13 | ND | 3.86 | 0.92 |
| PD | M | 65-69 | 94.40 | 44.32 | 19.48 | 1.01 | 10.03 | 3.31 | 10.40 | 3.72 |
| PD | F | 65-69 | 198.83 | 101.87 | ND | ND | ND | ND | ND | ND |
| PD | M | 90-94 | 101.25 | 55.55 | 23.52 | 3.46 | 9.60 | ND | 47.15 | 18.19 |

Bmax (fmol/mg) sites of ^3^H-JHU11761 (CSF1R) in frozen sections. GM, grey matter; WM, white matter.

Supplementary table 2.

| Region | F | p |  |  | t | p |  |
| --- | --- | --- | --- | --- | --- | --- | --- |
| Anterior cingulate | 1.65 | 0.217 |  | HC vs. Mild PD | -2.56 | 0.338 |  |
|  |  |  |  | HC vs. Mod PD | 1.13 | 0.866 |  |
|  |  |  |  | Mild PD vs. Mod PD | 3.69 | 0.263 |  |
| Brainstem | 5.25 | 0.015 | * | HC vs. Mild PD | -1.4 | 0.522 |  |
|  |  |  |  | HC vs. Mod PD | 3.88 | 0.060 |  |
|  |  |  |  | Mild PD vs. Mod PD | 5.28 | 0.012 | * |
| Cerebellar Cortex | 5.59 | 0.012 | * | HC vs. Mild PD | -1.02 | 0.738 |  |
|  |  |  |  | HC vs. Mod PD | 4.74 | 0.031 | * |
|  |  |  |  | Mild PD vs. Mod PD | 5.77 | 0.011 | * |
| Striatum | 9.68 | 0.001 | ** | HC vs. Mild PD | -0.68 | 0.902 |  |
|  |  |  |  | HC vs. Mod PD | 7.68 | 0.002 | ** |
|  |  |  |  | Mild PD vs. Mod PD | 8.35 | 0.002 | ** |
| Frontal cortex | 5.36 | 0.014 | * | HC vs. Mild PD | -1.13 | 0.646 |  |
|  |  |  |  | HC vs. Mod PD | 4.11 | 0.043 | * |
|  |  |  |  | Mild PD vs. Mod PD | 5.24 | 0.012 | * |
| Hippocampus | 6.39 | 0.008 | * | HC vs. Mild PD | -1.47 | 0.519 |  |
|  |  |  |  | HC vs. Mod PD | 4.59 | 0.031 | * |
|  |  |  |  | Mild PD vs. Mod PD | 6.05 | 0.006 | * |
| Occipital cortex | 3.24 | 0.062 |  | HC vs. Mild PD | -1.22 | 0.677 |  |
|  |  |  |  | HC vs. Mod PD | 3.46 | 0.156 |  |
|  |  |  |  | Mild PD vs. Mod PD | 4.68 | 0.051 |  |
| Parietal cortex | 5.75 | 0.011 | * | HC vs. Mild PD | -0.99 | 0.748 |  |
|  |  |  |  | HC vs. Mod PD | 4.81 | 0.027 | * |
|  |  |  |  | Mild PD vs. Mod PD | 5.80 | 0.010 | * |
| Posterior cingulate | 2.69 | 0.094 |  | HC vs. Mild PD | -2.08 | 0.477 |  |
|  |  |  |  | HC vs. Mod PD | 3.15 | 0.341 |  |
|  |  |  |  | Mild PD vs. Mod PD | 5.23 | 0.079 |  |
| Pallidum | 4.39 | 0.027 | * | HC vs. Mild PD | 1.48 | 0.716 |  |
|  |  |  |  | HC vs. Mod PD | 5.60 | 0.068 |  |
|  |  |  |  | Mild PD vs. Mod PD | 7.08 | 0.023 | * |
| Temporal cortex | 5.58 | 0.012 | * | HC vs. Mild PD | -1.86 | 0.457 |  |
|  |  |  |  | HC vs. Mod PD | 4.69 | 0.057 |  |
|  |  |  |  | Mild PD vs. Mod PD | 6.55 | 0.009 | * |
| Thalamus | 6.48 | 0.007 | * | HC vs. Mild PD | -1.13 | 0.713 |  |
|  |  |  |  | HC vs. Mod PD | 5.36 | 0.019 | * |
|  |  |  |  | Mild PD vs. Mod PD | 6.49 | 0.006 | * |

F values for ANOVA comparing regional [^11^C]CPPC V_T_ across healthy controls (HC), PD patients with mean MDS-UPDRS part II below median (Mild PD), and PD patients with mean MDS-UPDRS part II at median or above (Moderate PD), all with Hoehn & Yahr stage ≤2. T values for pair-wise post hoc tests between groups with Tukey’s test. *p<0.05 (uncorrected). Ϯp<0.005 (corrected for multiple comparisons).

Supplementary Figure 1.

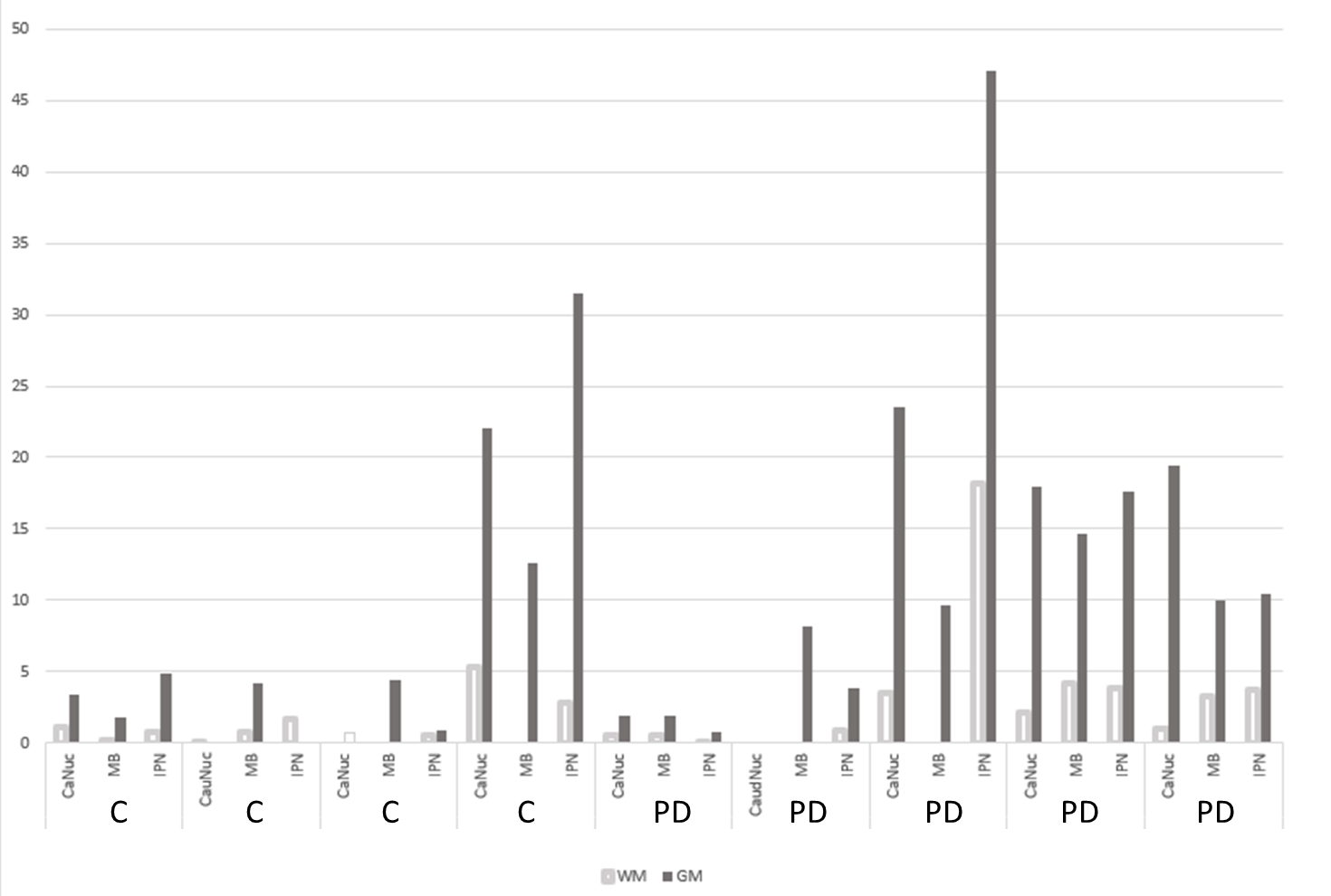

5 nM binding survey of ^3^H-JHU11761 binding in 5 healthy controls (C) and 4 subjects with Parkinson’s disease (PD) in inferior parietal cortex (IPC), midbrain (MB), and caudate nucleus (CaNuc).
